# Urinary Cell Gene Signature of Acute Rejection in Kidney Allografts

**DOI:** 10.1101/2023.12.18.23300165

**Authors:** Thalia Salinas, Carol Li, Catherine Snopkowski, Vijay K. Sharma, Darshana M. Dadhania, Karsten Suhre, Thangamani Muthukumar, Manikkam Suthanthiran

**Affiliations:** Division of Nephrology and Hypertension, Department of Medicine, Weill Cornell Medical College, New York, NY, USA; Department of Transplantation Medicine, New York-Presbyterian/Weill Cornell Medical Center, New York, NY, USA; Department of Physiology and Biophysics, Weill Cornell Medical College in Qatar, Doha, Qatar

**Author notes:** **Corresponding Author:** Thalia Salinas, MD, Division of Nephrology and Hypertension, Department of Transplantation Medicine, Weill Cornell Medical College, Address: 1300 York Avenue, 5^th^ floor, A-549, New York, NY, USA 10065.

**Keywords:** Allograft, Biomarker, Kidney, mRNA, PCR, Rejection

## Abstract

**Introduction:** A kidney allograft biopsy may display acute T cell-mediated rejection (TCMR), antibody-mediated rejection (ABMR), or concurrent TCMR + ABMR (MR). Development of noninvasive biomarkers diagnostic of all three types of acute rejection is a useful addition to the diagnostic armamentarium.

**Methods:** We developed customized RT-qPCR assays and measured urinary cell mRNA copy number in 145 biopsy-matched urine samples from 126 kidney allograft recipients and calculated urinary cell three-gene signature score from log_10_-transformed values for the 18S-normalized CD3E mRNA, 18S-normalized CXCL10 mRNA and 18S rRNA. We determined whether the signature score in biopsy-matched urine specimens discriminates biopsies without rejection (NR, n=50) from biopsies displaying TCMR (n=47), ABMR (n=28) or MR (n=20).

**Results:** Urinary cell three-gene signature discriminated TCMR, ABMR or MR biopsies from NR biopsies (P <0.0001, One-way ANOVA). Dunnett’s multiple comparisons test yielded P<0.0001 for NR vs. TCMR; P <0.001 for NR vs. ABMR; and P <0.0001 for NR vs. MR. By bootstrap resampling, optimism-corrected area under the receiver operating characteristic curve (AUC) was 0.749 (bias-corrected 95% confidence interval [CI], 0.638 to 0.840) for NR vs. TCMR (P<0.0001); 0.780 (95% CI, 0.656 to 0.878) for NR vs. ABMR (P<0.0001); and 0.857 (95% CI, 0.727 to 0.947) for NR vs. MR (P<0.0001). All three rejection biopsy categories were distinguished from NR biopsies with similar accuracy (all AUC comparisons P>0.05).

**Conclusion:** Urinary cell three-gene signature score may serve as a universal diagnostic signature of acute rejection due to TCMR, ABMR or MR in human kidney allografts with similar performance characteristics.

## INTRODUCTION

Kidney transplantation is the treatment of choice for patients with end-stage kidney disease. Unfortunately, the allograft recipient’s immune system perceives the life-saving allograft as non-self and mounts an allograft destructive immunity termed acute rejection.^1–3^ Acute rejection (AR) is categorized as T cell-mediated rejection (TCMR), antibody-mediated rejection (ABMR), or concurrent TCMR+ABMR (Mixed Rejection [MR]) based on histological features observed in the kidney allograft biopsy specimen.^4^ All three types of AR have been associated with graft dysfunction and suboptimal graft and patient survival rates.^5–9^

Percutaneous core needle biopsy is the method of choice for diagnosing AR in kidney allografts. The invasive biopsy procedure has been associated with bleeding, graft loss and even death. A recent systemic analysis concluded that true rates of these complications are not well defined because of the quality of existing data and possible publication bias.^10^ Also, the kidney is endowed with one million nephrons and at least two cores may be needed for accurate diagnosis due to sampling error.^11^ Inter-observer variability in biopsy interpretation^12^ and the logistics of capturing the dynamic alloimmune response by repeat biopsies represent additional challenges. Development of noninvasive biomarkers diagnostic of AR is therefore a high priority in the transplantation arena.

We developed urinary cell mRNA profiling for the noninvasive diagnosis of TCMR in human kidney allografts.^13–17^ In thematically linked urinary cell mRNA profiling studies, we identified higher abundance of mRNAs encoding cytotoxic attack molecules and mRNAs for immunoregulatory proteins in urine matched to TCMR biopsies compared to urine matched to biopsies without acute or chronic rejection features. Our single-center investigations led to a multicenter Clinical Trials in Organ Transplantation-04 (CTOT-04) study in which we measured absolute levels of mRNAs in 4300 urine specimens prospectively collected from 485 kidney allograft recipients.^18^ In the CTOT-04 study, we measured mRNAs we previously identified in our single-center studies to be associated with TCMR^13–17^ and developed and validated a parsimonious three-gene signature consisting of urinary cell levels of CD3E mRNA, IP-10 (CXCL10) mRNA and 18S rRNA (CTOT-04 signature) diagnostic and prognostic of TCMR.^18^ In the CTOT-04 study, there were only 10 ABMR biopsy-matched urine samples from 9 kidney allograft recipients, and we did not analyze whether the signature is also diagnostic of ABMR given the limited sample size.

A kidney allograft biopsy may display TCMR, ABMR, or MR. A noninvasive biomarker diagnostic of all three types of AR represents a significant advance and an unmet need in kidney transplantation. To fill the existing gap in knowledge, we investigated the hypothesis that the CTOT-04 signature is not only diagnostic of TCMR but also of ABMR and MR in kidney allografts. Our postulate was stimulated by our RNA sequencing study of kidney allografts and urinary cells that revealed shared gene expression patterns between urine matched to TCMR biopsies and urine matched to ABMR biopsies.^19^ Importantly, mRNAs for CD3E and CXCL10, the two mRNAs included in the CTOT-04 signature, were among the enriched and shared mRNAs in urine matched to TCMR biopsies vs. No Rejection biopsies and urine matched to ABMR biopsies vs. No Rejection biopsies.^19^

Herein, we report, using an independent cohort of kidney allograft recipients, that the CTOT-04 signature is not only diagnostic of TCMR but also of ABMR or MR in kidney allografts and the signature discriminates patients without rejection in their biopsies from those with acute rejection due to TCMR, ABMR or MR.

Kidney allograft recipients with TCMR, ABMR or MR often present with allograft dysfunction, and we examined whether the CTOT-04 signature is associated with allograft function. We considered it important to determine this since in our metabolomic profiling of biopsy-matched cell-free urine supernatants from kidney allograft recipients we found that estimated GFR (eGFR) is a confounder of metabolite-based signatures of AR.^20^ We report here the clinically relevant finding that eGFR is not associated with the CTOT-04 signature score and is accurately diagnostic of AR after controlling for eGFR.

## MATERIALS AND METHODS

### Kidney allograft recipients

We measured urinary cell levels of CD3E mRNA, IP-10 (CXCL10) mRNA and 18S rRNA in 145 urine samples matched to 145 kidney allograft biopsies from 126 unique kidney allograft recipients. The study was approved by the Weill Cornell Medicine Institutional Review Board and study participants provided written informed consent for participation. The investigation was performed in compliance with the ethical principles specified in the Declaration of Helsinki.^21–22^

### Kidney allograft biopsies

Percutaneous needle core biopsies were performed under ultrasound guidance at the NewYork Presbyterian Hospital/Weill Cornell Medicine. Biopsies were fixed and stained using hematoxylin eosin, periodic acid Schiff and Masson trichrome stains. Biopsies were classified and graded by our transplant pathologists (S.S and S.V.S) using the Banff 2019 update of the Banff ‘97 classification of allograft pathology.^4^

Biopsies were stained for complement component 4 degradation product (C4d) and Simian Vacuolating virus large T antigen (SV40 large T antigen). Intragraft presence or absence of C4d was determined by immunofluorescence microscopy of frozen section. The presence or absence of SV40 large T antigen was determined by immunostaining with mouse monoclonal IgG2a antibody PAb416 reacting with both SV40 antigen and BKV large T antigen.

### Urine biospecimens, isolation of RNA and reverse transcription

Approximately 50 mL of urine was obtained from each recipient using a sterile urine cup at the time of biopsy and was processed for mRNA profiling at our Gene Expression Monitoring (GEM) Core at Weill Cornell Medicine, New York, NY. Urine samples were centrifuged at 2000*g* for 30 minutes at room temperature to prepare urinary cell pellets. RNalater (50µl), RLT buffer and Beta-mercaptoethanol (350 μl) were added to the sedimented cells and lysed. Total RNA was isolated from the cell lysate using the RNeasy mini kit (Qiagen, Cat. 74104). RNA purity (ratio of A260/A280) and yield (absorbance at A260) were measured using NanoDrop ND-1000 spectrophotometer (ThermoFisher Scientific).

Total RNA was reverse transcribed (RT) to cDNA using TaqMan reverse transcription kit (Cat. N808-0234, Applied Biosystems). RT was performed at the total RNA concentration of 1.0 μg in 100 μl volume and the RT reaction contained 1x TaqMan RT buffer, 500 μM each of 4 dNTPs, 2.5 μM of Random Hexamer, 0.4 Unit/μl of RNase inhibitor, 1.25 Unit/μl of MultiScribe Reverse Transcriptase and 5.5 mM of Magnesium Chloride. The RT mixture was incubated at 25°C for 10 min, 48°C for 30 min, and 95°C for 5 min.

### Design and development of gene-specific oligonucleotide primer pairs and TaqMan probes

We designed gene-specific oligonucleotide primers and TaqMan fluorogenic probes (hydrolysis probes) for the measurement of mRNA levels for CD3E, IP-10, TGF-β1 and 18S rRNA; TGF-β1 mRNA was measured as a QC parameter of customized RT-qPCR assays and 18S rRNA served both as a component of the CTOT-04 signature and as a QC parameter. The sequence and location of the gene-specific oligonucleotide primer pairs and TaqMan probes utilized have been reported.^18^

### Preamplification of cDNA with gene-specific primer pairs

We developed the preamplification protocol to compensate for low yield of total RNA from urinary cells and to measure multiple mRNAs from small amounts of cDNA.^18^ The preamplification reaction for each sample was set up to a final reaction volume of 10.0 μl containing 3.0 μl cDNA (from RT of 1 μg total RNA in 100 μl buffer), 5.0 µl Platinum® Multiplex PCR Master Mix, 1.68 µl primer mix (50 μM sense and 50 μM antisense primer) and 0.32 μl water to a final volume of 10 μl. The PCR was set up in a Veriti thermal cycler (Applied Biosystems) and the reaction profile consisted of an initial hold at 95°C for 2 min, 11 cycles of denaturing at 95°C for 30 seconds, primer annealing at 60°C for 90 seconds, extension at 72°C for 1 min, and final hold at 72°C for 10 min. At the end of 11 cycles of amplification, 290 μl of TE buffer was added to the PCR reaction, and 2.5 μl of diluted PCR amplicons were then used for quantification of mRNA using the RT-qPCR assay.

### Absolute Quantification of mRNA in customized RT-qPCR assay

We quantified absolute levels of mRNAs using in-house synthesized BAK amplicon in the customized RT-qPCR assay. The Bak amplicon was diluted to a concentration of 1E+7copies/μl (stock solution) and the stock solution was diluted over 6 orders of magnitude (1,000,000, 100,000, 10,000, 1,000, 100, and 10 copies per 1 μl) (work solution) to develop the universal standard curve. The standard curve was developed by adding 2.5 μl of diluted stock solution to duplicate wells and amplified with Bak-specific primer pair and Bak-specific fluorogenic TaqMan probe. The threshold cycles (C_T_) were plotted against the log of the initial amount of the Bak amplicon to develop the standard curve. The standard curve copy numbers in our RT-qPCR assays ranged from 25 to 2.5 million copies.

### Calculation of CTOT-04 signature

CTOT-04 signature in urinary cells was calculated using the previously reported equation: – 6.1487+0.8534log_10_(CD3E/18S) + 0.6376log_10_(IP-10/18S) +1.6464log_10_(18S).^18^

### Data analysis

Log_10_ transformation of mRNA copy number reduced deviation of mRNA copy number from the normal distribution. mRNA copy number in each urine sample was normalized by calculating the ratio of mRNA copy number to copy number of the reference gene 18S rRNA in the same urine sample. mRNA copy numbers <25 (including 0) were scored as 12.5 copies per microgram of total RNA before 18S rRNA normalization. T test was used for two group comparisons, one-way Analysis of Variance (ANOVA) for multiple group comparison and Dunnett’s test following ANOVA for pair-wise comparisons. The ability of the CTOT-04 signature to discriminate between Banff biopsy diagnostic categories was evaluated using receiver operating characteristic (ROC) curves and area under the ROC curve (AUC) and 95% confidence intervals (95% CI) were calculated. For robust statistical inference, we generated 5000 bootstrap sample sets and estimated the optimism-corrected AUCs of ROC curves and bias-corrected 95% CI. Sensitivity and specificity were determined using the previously identified CTOT-04 signature cutpoint of -1.213, the Youden index that maximized sensitivity and specificity for discrimination of those with TCMR biopsies from those with No Rejection (NR) biopsies.^18^ The associations of the CTOT-04 signature with the covariates eGFR and time since transplantation to biopsy were analyzed using the R function lm() from the “stats” package (R version 4.2.1) in univariate and multivariate models,^23^ and associations with the biopsy diagnosis were computed using the R function glm(), e.g., glm(as.factor(NRvs.AR)∼CTOTsig+eGFR,=“binomial”).^24^ The corresponding summary statistics were obtained using the corresponding R summary.

## RESULTS

### Characteristics of Kidney allograft recipients and biopsy-related parameters

The study cohort consisted of 126 unique kidney allograft recipients (patients) and 145 kidney allograft biopsy-matched urine specimens. The median (25^th^ percentile and 75^th^ percentile) age was 48.3 years (36.0 and 48.3) and there were 56 females and 70 males. The eGFR, calculated using the 2021 CKD-EPI equation without using race^25^ was 27 ml (16 and 30), and time since transplantation to biopsy was 7.3 months (1.8 and 39.8). **Table 1** shows patient characteristics stratified by biopsy diagnosis. Biopsies were selected from our Biorepository to represent all three major types of AR. As reflected by the higher number of biopsies (n=145) compared to patients (n=126), 17 patients contributed more than one biopsy, and among those biopsies from 6 patients belonged to more than one diagnostic category. As the control group for patients with AR biopsies, we selected patients with biopsies with no rejection features and varying degrees of tubular injury (NR).

**Table 1.**
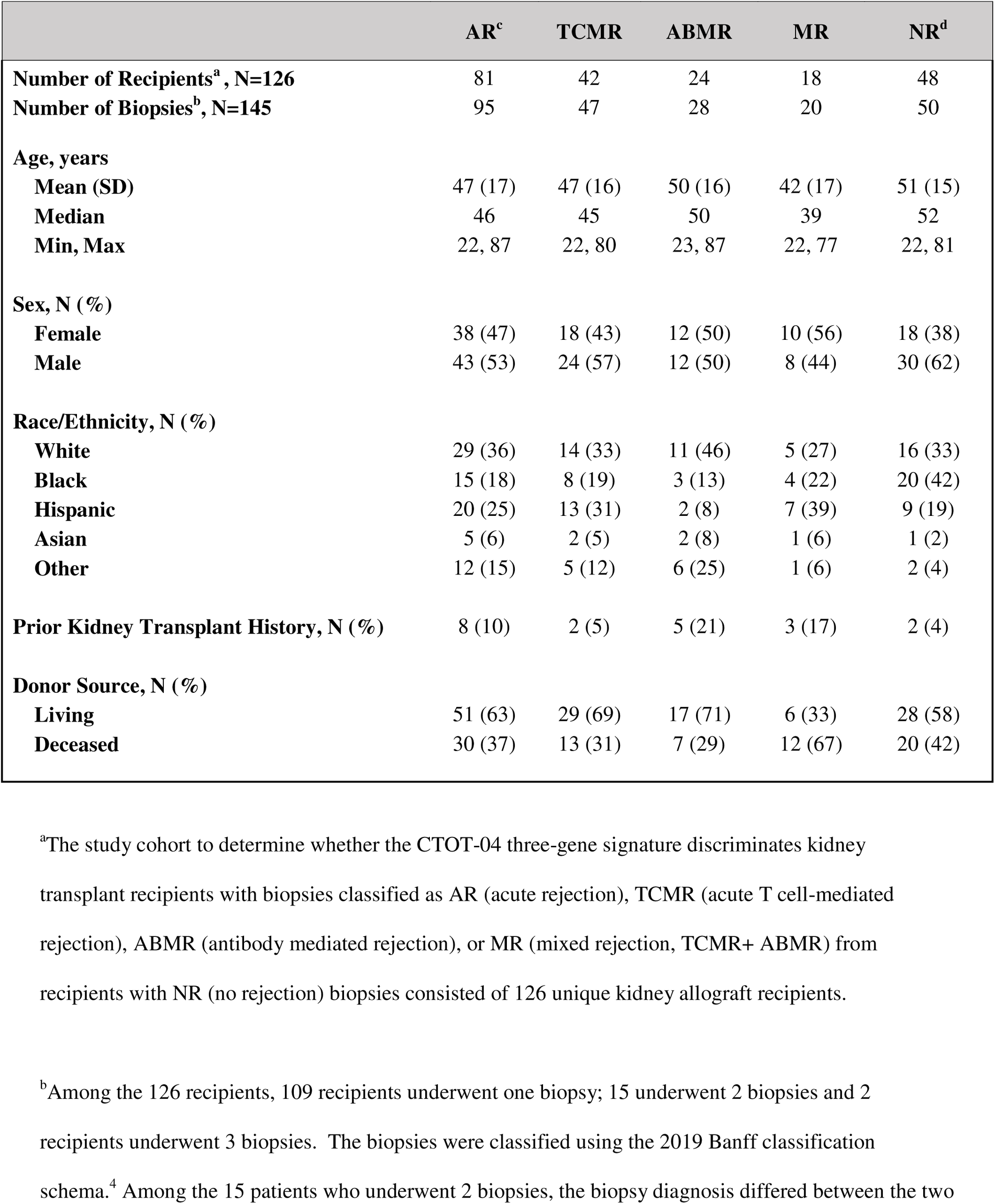
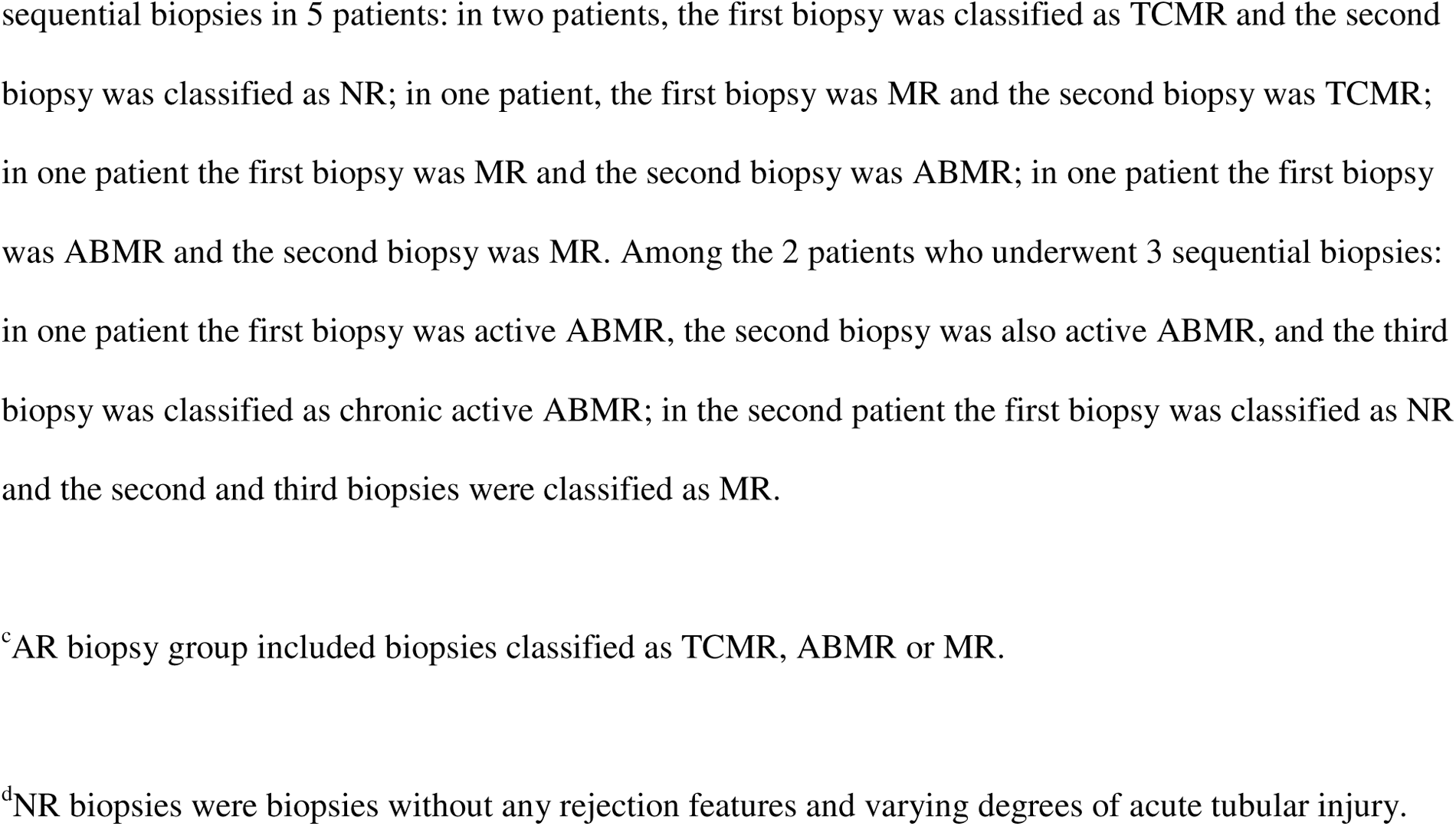
Characteristics of Kidney Transplant Recipients.

|  | AR<sup>c</sup> | TCMR | ABMR | MR | NR<sup>d</sup> |
| --- | --- | --- | --- | --- | --- |
| **Number of Recipients<sup>a</sup>, N=126** | 81 | 42 | 24 | 18 | 48 |
| **Number of Biopsies<sup>b</sup>, N=145** | 95 | 47 | 28 | 20 | 50 |
| **Age, years** |  |  |  |  |  |
| **Mean (SD)** | 47 (17) | 47 (16) | 50 (16) | 42 (17) | 51 (15) |
| **Median** | 46 | 45 | 50 | 39 | 52 |
| **Min, Max** | 22, 87 | 22, 80 | 23, 87 | 22, 77 | 22, 81 |
| **Sex, N (%)** |  |  |  |  |  |
| **Female** | 38 (47) | 18 (43) | 12 (50) | 10 (56) | 18 (38) |
| **Male** | 43 (53) | 24 (57) | 12 (50) | 8 (44) | 30 (62) |
| **Race/Ethnicity, N (%)** |  |  |  |  |  |
| **White** | 29 (36) | 14 (33) | 11 (46) | 5 (27) | 16 (33) |
| **Black** | 15 (18) | 8 (19) | 3 (13) | 4 (22) | 20 (42) |
| **Hispanic** | 20 (25) | 13 (31) | 2 (8) | 7 (39) | 9 (19) |
| **Asian** | 5 (6) | 2 (5) | 2 (8) | 1 (6) | 1 (2) |
| **Other** | 12 (15) | 5 (12) | 6 (25) | 1 (6) | 2 (4) |
| **Prior Kidney Transplant History, N (%)** | 8 (10) | 2 (5) | 5 (21) | 3 (17) | 2 (4) |
| **Donor Source, N (%)** |  |  |  |  |  |
| **Living** | 51 (63) | 29 (69) | 17 (71) | 6 (33) | 28 (58) |
| **Deceased** | 30 (37) | 13 (31) | 7 (29) | 12 (67) | 20 (42) |

<sup>a</sup>The study cohort to determine whether the CTOT-04 three-gene signature discriminates kidney transplant recipients with biopsies classified as AR (acute rejection), TCMR (acute T cell-mediated rejection), ABMR (antibody mediated rejection), or MR (mixed rejection, TCMR+ ABMR) from recipients with NR (no rejection) biopsies consisted of 126 unique kidney allograft recipients.
<sup>b</sup>Among the 126 recipients, 109 recipients underwent one biopsy; 15 underwent 2 biopsies and 2 recipients underwent 3 biopsies. The biopsies were classified using the 2019 Banff classification schema.<sup>4</sup> Among the 15 patients who underwent 2 biopsies, the biopsy diagnosis differed between the two
sequential biopsies in 5 patients: in two patients, the first biopsy was classified as TCMR and the second biopsy was classified as NR; in one patient, the first biopsy was MR and the second biopsy was TCMR; in one patient the first biopsy was MR and the second biopsy was ABMR; in one patient the first biopsy was ABMR and the second biopsy was MR. Among the 2 patients who underwent 3 sequential biopsies: in one patient the first biopsy was active ABMR, the second biopsy was also active ABMR, and the third biopsy was classified as chronic active ABMR; in the second patient the first biopsy was classified as NR and the second and third biopsies were classified as MR.
<sup>c</sup>AR biopsy group included biopsies classified as TCMR, ABMR or MR.
<sup>d</sup>NR biopsies were biopsies without any rejection features and varying degrees of acute tubular injury.

Kidney allograft biopsy-related parameters, stratified by biopsy diagnosis, are shown (**Table 2**). Among the 145 biopsies, 138 were clinically indicated (for-cause) biopsies. All biopsies were judged adequate by pathologists and the mean (SD) number of glomeruli in each diagnostic category met or exceeded Banff thresholds for adequacy (**Table 2**). Ninety-five of the 145 biopsies were grouped as AR biopsies with 47 biopsies from 42 patients categorized as TCMR; 28 biopsies from 24 patients classified as ABMR; and 20 biopsies from 18 patients classified as MR. Fifty biopsies from 48 patients were classified as NR. All biopsies were negative for intragraft SV40 large T antigen.

**Table 2.**
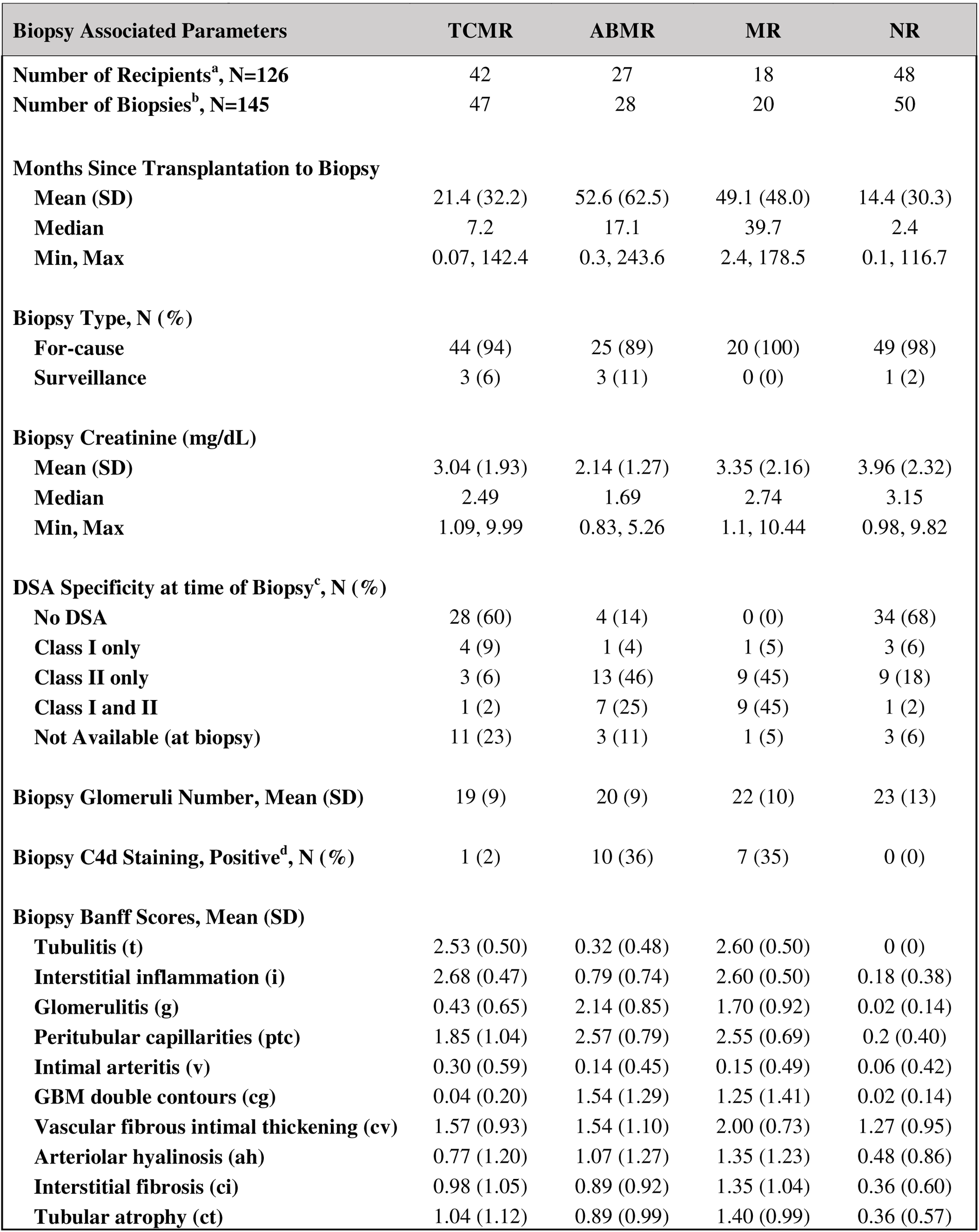
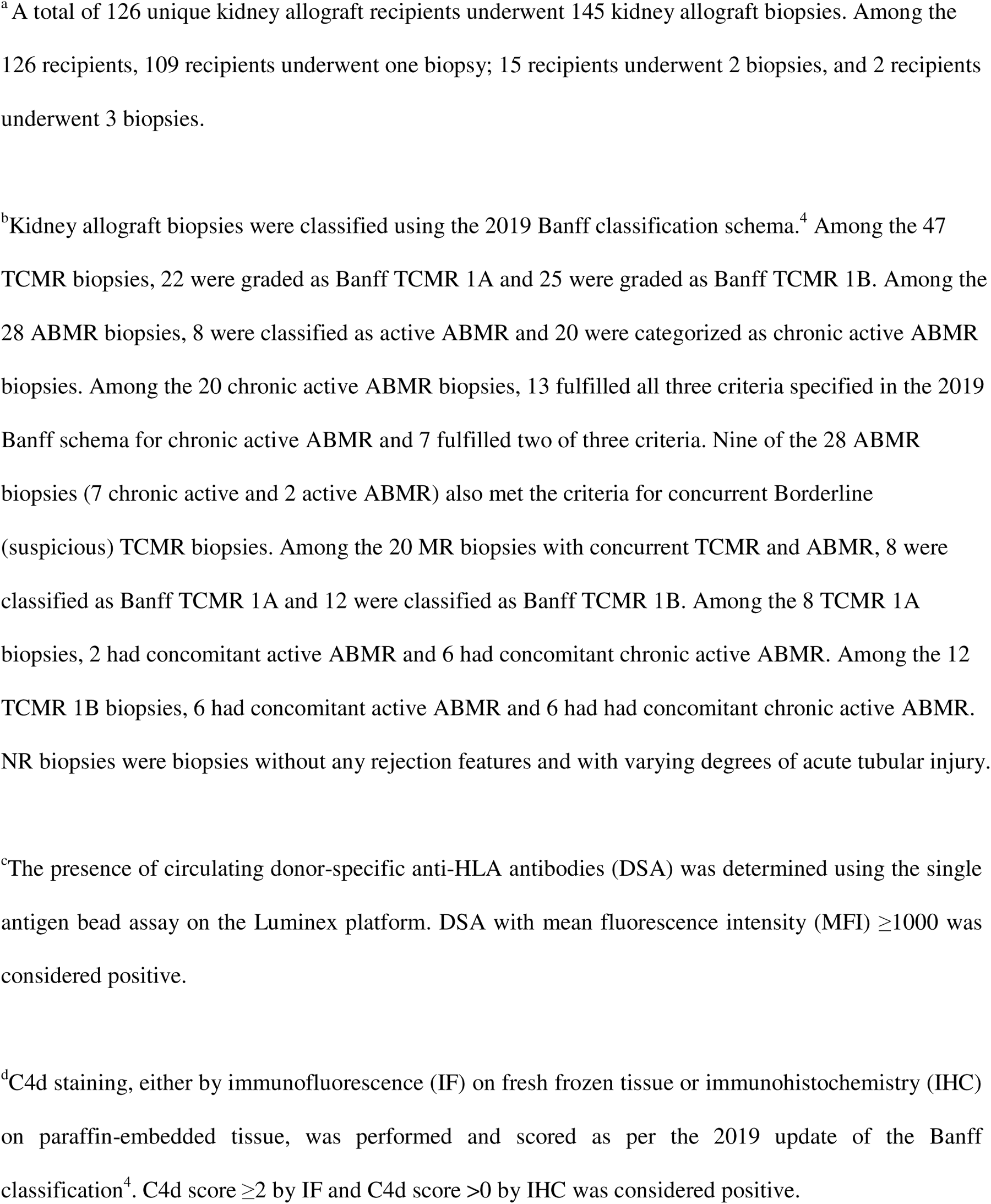
Kidney Allograft Biopsy Associated Parameters and Banff Lesion Scores.

| **Biopsy Associated Parameters** | **TCMR** | **ABMR** | **MR** | **NR** |
| --- | --- | --- | --- | --- |
| **Number of Recipients<sup>a</sup>, N=126** | 42 | 27 | 18 | 48 |
| **Number of Biopsies<sup>b</sup>, N=145** | 47 | 28 | 20 | 50 |
| **Months Since Transplantation to Biopsy** |  |  |  |  |
| **Mean (SD)** | 21.4 (32.2) | 52.6 (62.5) | 49.1 (48.0) | 14.4 (30.3) |
| **Median** | 7.2 | 17.1 | 39.7 | 2.4 |
| **Min, Max** | 0.07, 142.4 | 0.3, 243.6 | 2.4, 178.5 | 0.1, 116.7 |
| **Biopsy Type, N (%)** |  |  |  |  |
| **For-cause** | 44 (94) | 25 (89) | 20 (100) | 49 (98) |
| **Surveillance** | 3 (6) | 3 (11) | 0 (0) | 1 (2) |
| **Biopsy Creatinine (mg/dL)** |  |  |  |  |
| **Mean (SD)** | 3.04 (1.93) | 2.14 (1.27) | 3.35 (2.16) | 3.96 (2.32) |
| **Median** | 2.49 | 1.69 | 2.74 | 3.15 |
| **Min, Max** | 1.09, 9.99 | 0.83, 5.26 | 1.1, 10.44 | 0.98, 9.82 |
| **DSA Specificity at time of Biopsy<sup>c</sup>, N (%)** |  |  |  |  |
| **No DSA** | 28 (60) | 4 (14) | 0 (0) | 34 (68) |
| **Class I only** | 4 (9) | 1 (4) | 1 (5) | 3 (6) |
| **Class II only** | 3 (6) | 13 (46) | 9 (45) | 9 (18) |
| **Class I and II** | 1 (2) | 7 (25) | 9 (45) | 1 (2) |
| **Not Available (at biopsy)** | 11 (23) | 3 (11) | 1 (5) | 3 (6) |
| **Biopsy Glomeruli Number, Mean (SD)** | 19 (9) | 20 (9) | 22 (10) | 23 (13) |
| **Biopsy C4d Staining, Positive<sup>d</sup>, N (%)** | 1 (2) | 10 (36) | 7 (35) | 0 (0) |
| **Biopsy Banff Scores, Mean (SD)** |  |  |  |  |
| **Tubulitis (t)** | 2.53 (0.50) | 0.32 (0.48) | 2.60 (0.50) | 0 (0) |
| **Interstitial inflammation (i)** | 2.68 (0.47) | 0.79 (0.74) | 2.60 (0.50) | 0.18 (0.38) |
| **Glomerulitis (g)** | 0.43 (0.65) | 2.14 (0.85) | 1.70 (0.92) | 0.02 (0.14) |
| **Peritubular capillarities (ptc)** | 1.85 (1.04) | 2.57 (0.79) | 2.55 (0.69) | 0.2 (0.40) |
| **Intimal arteritis (v)** | 0.30 (0.59) | 0.14 (0.45) | 0.15 (0.49) | 0.06 (0.42) |
| **GBM double contours (cg)** | 0.04 (0.20) | 1.54 (1.29) | 1.25 (1.41) | 0.02 (0.14) |
| **Vascular fibrous intimal thickening (cv)** | 1.57 (0.93) | 1.54 (1.10) | 2.00 (0.73) | 1.27 (0.95) |
| **Arteriolar hyalinosis (ah)** | 0.77 (1.20) | 1.07 (1.27) | 1.35 (1.23) | 0.48 (0.86) |
| **Interstitial fibrosis (ci)** | 0.98 (1.05) | 0.89 (0.92) | 1.35 (1.04) | 0.36 (0.60) |
| **Tubular atrophy (ct)** | 1.04 (1.12) | 0.89 (0.99) | 1.40 (0.99) | 0.36 (0.57) |

<sup>a</sup> A total of 126 unique kidney allograft recipients underwent 145 kidney allograft biopsies. Among the 126 recipients, 109 recipients underwent one biopsy; 15 recipients underwent 2 biopsies, and 2 recipients underwent 3 biopsies.
<sup>b</sup> Kidney allograft biopsies were classified using the 2019 Banff classification schema.<sup>4</sup> Among the 47 TCMR biopsies, 22 were graded as Banff TCMR 1A and 25 were graded as Banff TCMR 1B. Among the 28 ABMR biopsies, 8 were classified as active ABMR and 20 were categorized as chronic active ABMR biopsies. Among the 20 chronic active ABMR biopsies, 13 fulfilled all three criteria specified in the 2019 Banff schema for chronic active ABMR and 7 fulfilled two of three criteria. Nine of the 28 ABMR biopsies (7 chronic active and 2 active ABMR) also met the criteria for concurrent Borderline (suspicious) TCMR biopsies. Among the 20 MR biopsies with concurrent TCMR and ABMR, 8 were classified as Banff TCMR 1A and 12 were classified as Banff TCMR 1B. Among the 8 TCMR 1A biopsies, 2 had concomitant active ABMR and 6 had concomitant chronic active ABMR. Among the 12 TCMR 1B biopsies, 6 had concomitant active ABMR and 6 had had concomitant chronic active ABMR. NR biopsies were biopsies without any rejection features and with varying degrees of acute tubular injury.
<sup>c</sup> The presence of circulating donor-specific anti-HLA antibodies (DSA) was determined using the single antigen bead assay on the Luminex platform. DSA with mean fluorescence intensity (MFI) $\geq 1000$ was considered positive.
<sup>d</sup> C4d staining, either by immunofluorescence (IF) on fresh frozen tissue or immunohistochemistry (IHC) on paraffin-embedded tissue, was performed and scored as per the 2019 update of the Banff classification<sup>4</sup>. C4d score $\geq 2$ by IF and C4d score $>0$ by IHC was considered positive.

### CTOT-04 signature score is diagnostic of acute rejection in kidney allografts

Figure 1 displays the CTOT-04 signature score in each biopsy-matched urine sample within the violin plots and the distribution of scores is represented by the density shape of the plot. The thin black lines within each plot are the 25^th^ and 75^th^ percentile values, and the thick black line is the median value of the CTOT-04 signature score. The black horizontal dotted line crossing the contours of both violin plots is the CTOT-04 signature score of -1.213 which maximized sensitivity and specificity for distinguishing TCMR biopsies from NR biopsies in the CTOT-04 study.^18^

**Figure 1.**
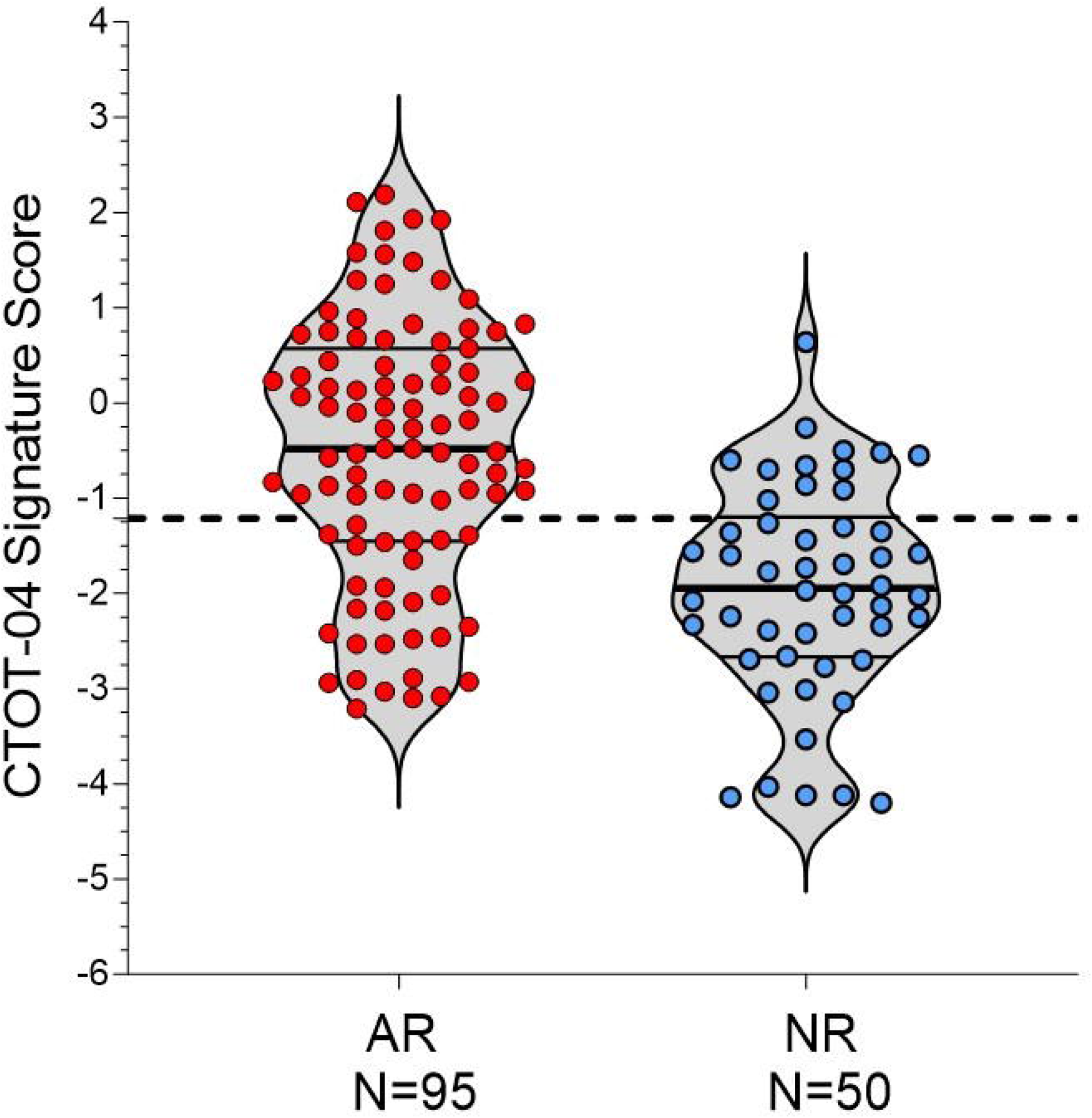
Violin Plots of the CTOT-04 Signature Scores in Urine matched to AR biopsies or NR biopsies. Violin plots of CTOT-04 signature scores in the urine samples matched to AR biopsies (N=95) or urine samples matched to NR biopsies (N=50). Customized RT-qPCR assays were used to measure urinary cell levels of CD3E mRNA, CXCL10 mRNA, and 18s rRNA in kidney allograft biopsy-matched urine samples collected from kidney allograft recipients and the CTOT-04 signature score was calculated from log10-transformed values for the 18S-normalized CD3E mRNA, 18S-normalized CXCL10 mRNA and 18S rRNA.18 The CTOT-04 signature score values of individual urine samples are shown as circles within the plot. The two thin black lines within each plot are the 25th and 75th percentile values, and thick black line is the 50th (median) value of the CTOT-04 signature score. The horizontal dotted line parallel to the x-axis is the previously identified CTOT-04 signature score cutpoint of -1.213 that maximized sensitivity and specificity for the diagnosis of TCMR in the multicenter CTOT-04 study of 485 prospectively enrolled kidney allograft recipients.^18^

In the current study, the median (25^th^ percentile and 75^th^ percentile) CTOT-04 signature score was -0.480 (-1.450 and 0.570) in the 95 urine samples matched to 95 AR biopsies and significantly higher than the score of -1.945 (-2.668 and -1.200) in the 50 urine samples matched to 50 NR biopsies (**Table 3**; P<0.0001, T test). The previously validated cutpoint of -1.213 for TCMR biopsy diagnosis distinguished urine samples matched to AR biopsies consisting of TCMR, ABMR and MR from urine samples matched to NR biopsies at a specificity of 76% and a sensitivity of 71% (**Table 4**; P<0.0001, Fischer’s Exact test).

**Table 3.**
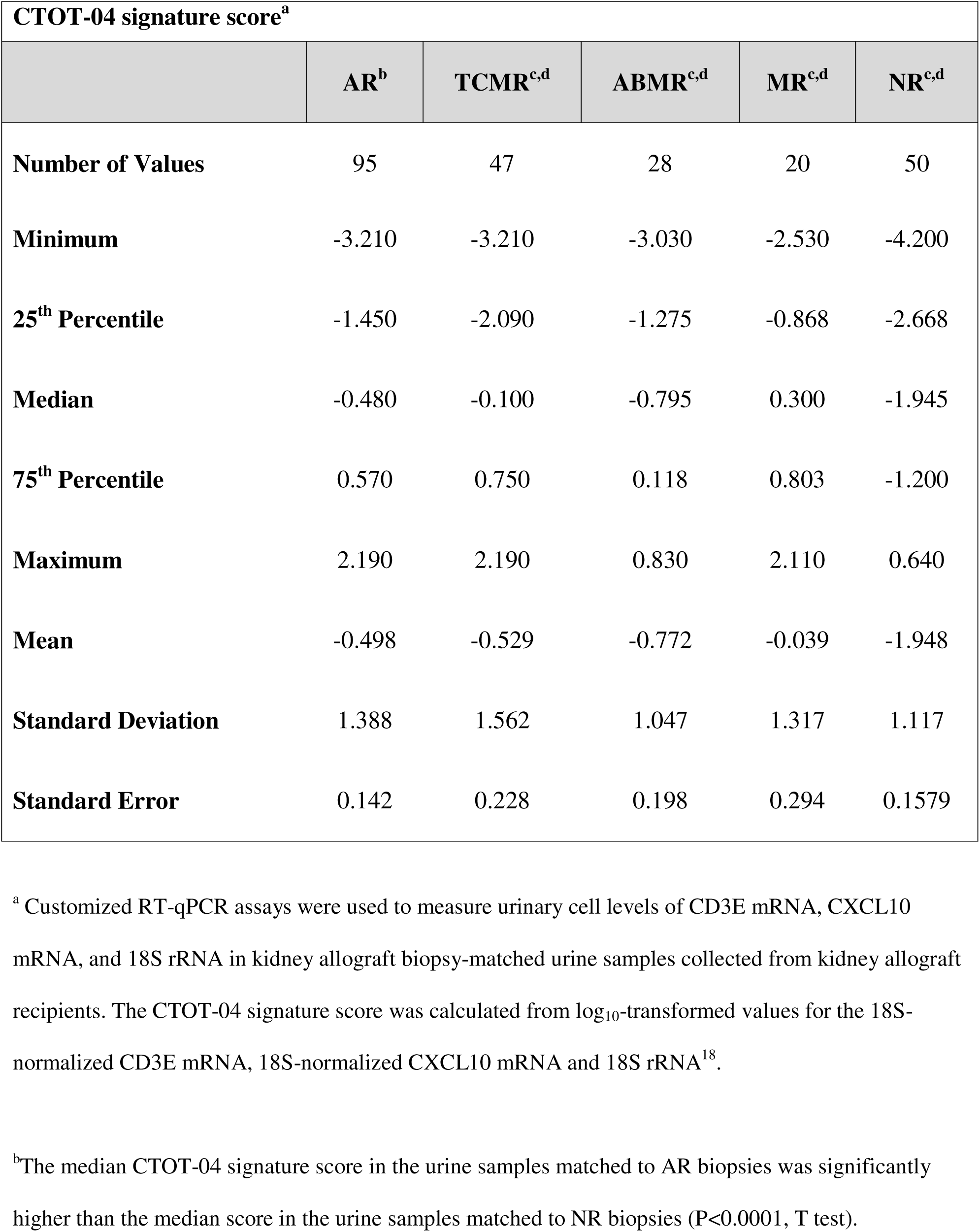
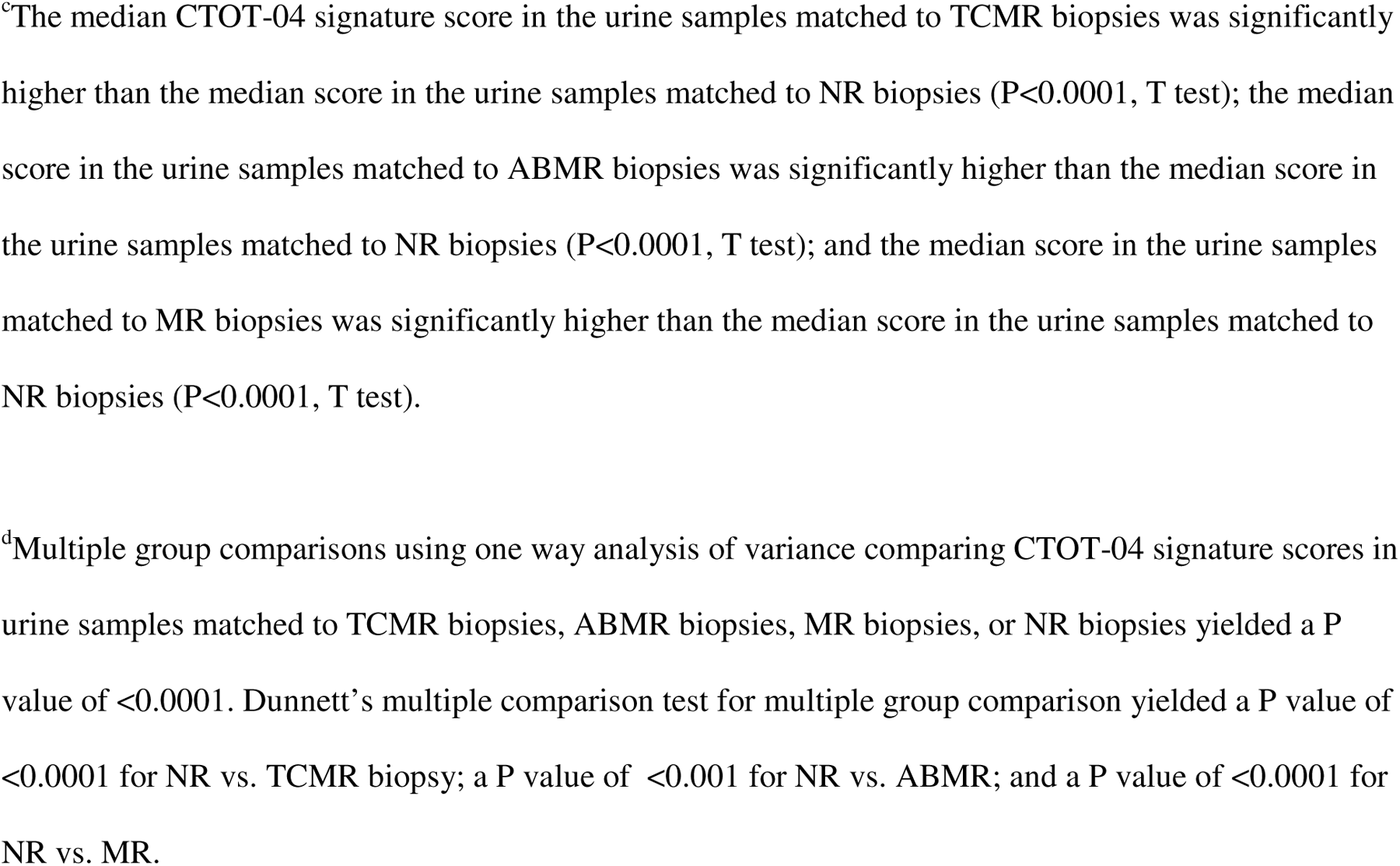
CTOT-04 Signature Scores in Urine Matched to Biopsies classified as AR, TCMR, ABMR, MR, or NR biopsies.

| **CTOT-04 signature score<sup>a</sup>** |  |  |  |  |  |
| --- | --- | --- | --- | --- | --- |
|  | **AR<sup>b</sup>** | **TCMR<sup>c,d</sup>** | **ABMR<sup>c,d</sup>** | **MR<sup>c,d</sup>** | **NR<sup>c,d</sup>** |
| **Number of Values** | 95 | 47 | 28 | 20 | 50 |
| **Minimum** | -3.210 | -3.210 | -3.030 | -2.530 | -4.200 |
| **25<sup>th</sup> Percentile** | -1.450 | -2.090 | -1.275 | -0.868 | -2.668 |
| **Median** | -0.480 | -0.100 | -0.795 | 0.300 | -1.945 |
| **75<sup>th</sup> Percentile** | 0.570 | 0.750 | 0.118 | 0.803 | -1.200 |
| **Maximum** | 2.190 | 2.190 | 0.830 | 2.110 | 0.640 |
| **Mean** | -0.498 | -0.529 | -0.772 | -0.039 | -1.948 |
| **Standard Deviation** | 1.388 | 1.562 | 1.047 | 1.317 | 1.117 |
| **Standard Error** | 0.142 | 0.228 | 0.198 | 0.294 | 0.1579 |

<sup>a</sup> Customized RT-qPCR assays were used to measure urinary cell levels of CD3E mRNA, CXCL10 mRNA, and 18S rRNA in kidney allograft biopsy-matched urine samples collected from kidney allograft recipients. The CTOT-04 signature score was calculated from log<sub>10</sub>-transformed values for the 18S-normalized CD3E mRNA, 18S-normalized CXCL10 mRNA and 18S rRNA<sup>18</sup>.
<sup>b</sup>The median CTOT-04 signature score in the urine samples matched to AR biopsies was significantly higher than the median score in the urine samples matched to NR biopsies (P<0.0001, T test).
<sup>c</sup>The median CTOT-04 signature score in the urine samples matched to TCMR biopsies was significantly higher than the median score in the urine samples matched to NR biopsies ( $P < 0.0001$ , T test); the median score in the urine samples matched to ABMR biopsies was significantly higher than the median score in the urine samples matched to NR biopsies ( $P < 0.0001$ , T test); and the median score in the urine samples matched to MR biopsies was significantly higher than the median score in the urine samples matched to NR biopsies ( $P < 0.0001$ , T test).
<sup>d</sup>Multiple group comparisons using one way analysis of variance comparing CTOT-04 signature scores in urine samples matched to TCMR biopsies, ABMR biopsies, MR biopsies, or NR biopsies yielded a P value of $< 0.0001$ . Dunnett's multiple comparison test for multiple group comparison yielded a P value of $< 0.0001$ for NR vs. TCMR biopsy; a P value of $< 0.001$ for NR vs. ABMR; and a P value of $< 0.0001$ for NR vs. MR.

**Table 4.**
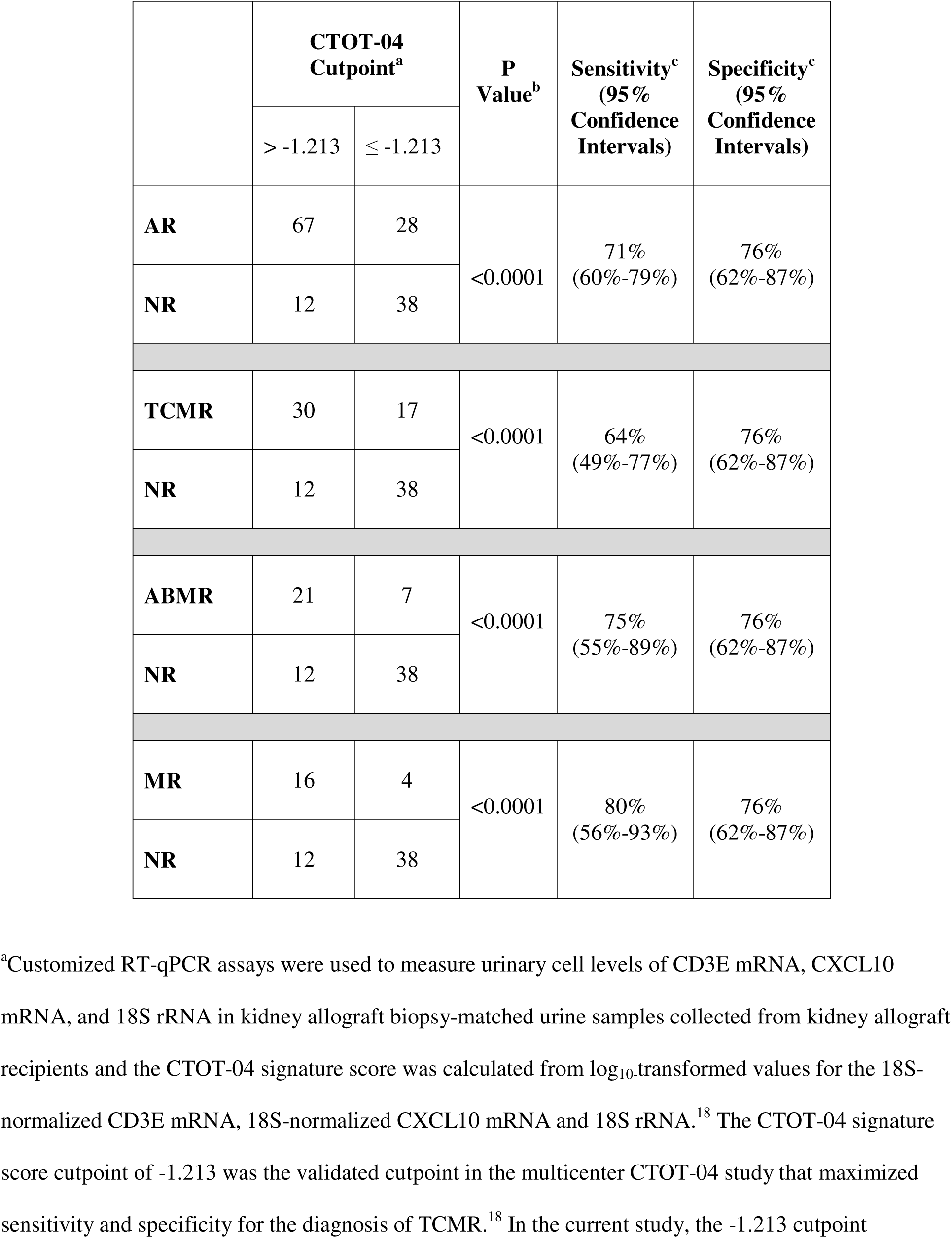
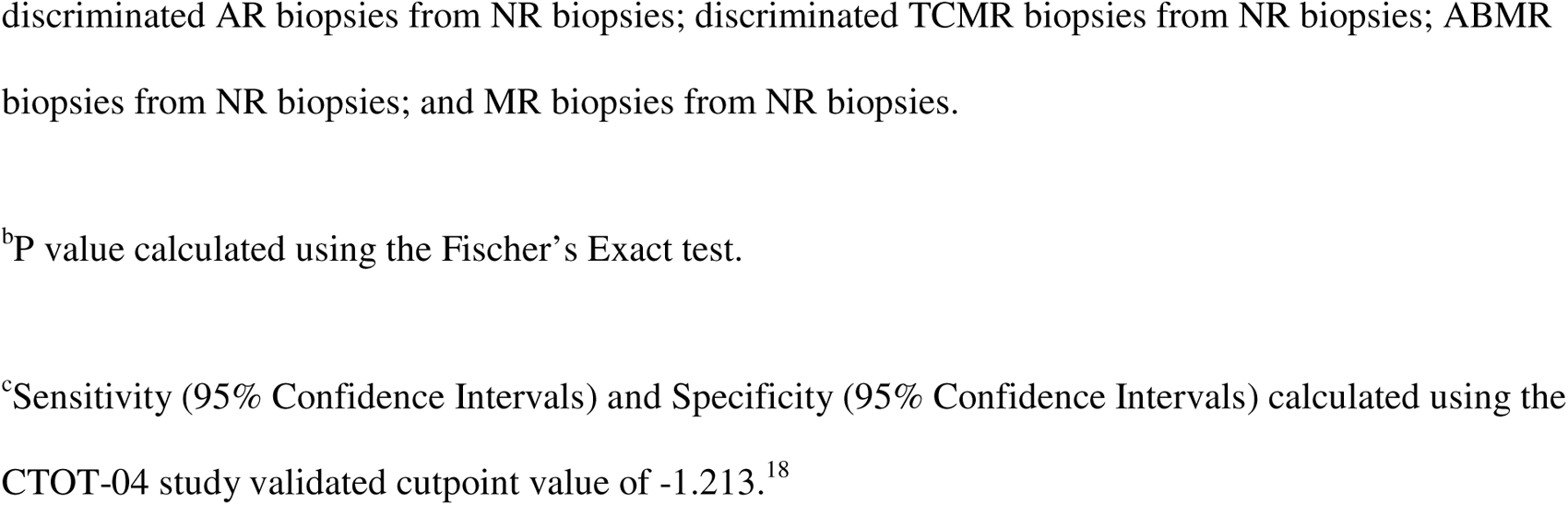
CTOT-04 signature score cutpoint of -1.213 discriminates kidney allograft recipients with AR, TCMR, ABMR, or MR biopsies from recipients with NR biopsies.

|  | CTOT-04<br>Cutpoint<sup>a</sup> |  | P<br>Value<sup>b</sup> | Sensitivity<sup>c</sup><br>(95%<br>Confidence<br>Intervals) | Specificity<sup>c</sup><br>(95%<br>Confidence<br>Intervals) |
| --- | --- | --- | --- | --- | --- |
|  | > -1.213 | ≤ -1.213 |  |  |  |
| AR | 67 | 28 | <0.0001 | 71%<br>(60%-79%) | 76%<br>(62%-87%) |
| NR | 12 | 38 |  |  |  |
| TCMR | 30 | 17 | <0.0001 | 64%<br>(49%-77%) | 76%<br>(62%-87%) |
| NR | 12 | 38 |  |  |  |
| ABMR | 21 | 7 | <0.0001 | 75%<br>(55%-89%) | 76%<br>(62%-87%) |
| NR | 12 | 38 |  |  |  |
| MR | 16 | 4 | <0.0001 | 80%<br>(56%-93%) | 76%<br>(62%-87%) |
| NR | 12 | 38 |  |  |  |

<sup>a</sup>Customized RT-qPCR assays were used to measure urinary cell levels of CD3E mRNA, CXCL10 mRNA, and 18S rRNA in kidney allograft biopsy-matched urine samples collected from kidney allograft recipients and the CTOT-04 signature score was calculated from log<sub>10</sub>-transformed values for the 18S-normalized CD3E mRNA, 18S-normalized CXCL10 mRNA and 18S rRNA.<sup>18</sup> The CTOT-04 signature score cutpoint of -1.213 was the validated cutpoint in the multicenter CTOT-04 study that maximized sensitivity and specificity for the diagnosis of TCMR.<sup>18</sup> In the current study, the -1.213 cutpoint
discriminated AR biopsies from NR biopsies; discriminated TCMR biopsies from NR biopsies; ABMR biopsies from NR biopsies; and MR biopsies from NR biopsies.
<sup>b</sup>P value calculated using the Fischer's Exact test.
<sup>c</sup>Sensitivity (95% Confidence Intervals) and Specificity (95% Confidence Intervals) calculated using the CTOT-04 study validated cutpoint value of -1.213.<sup>18</sup>

The median (25^th^ and 75^th^ percentiles) eGFR was 29 mL (18 and 38) in the AR group and 20 mL (11 and 33) in the NR group (P=0.0025). We examined whether the CTOT-04 signature score was associated with eGFR and determined that it was not associated (F-statistic: 0.3125, on 1 and 143 degrees of freedom, P=0.577).^23^ We also found that the CTOT-04 signature score was diagnostic of AR after controlling for eGFR: P=3.88e-07 for the association without accounting for eGFR compared to P=6.4e-07 including eGFR in a general linear model.^24^

The median time since transplantation to biopsy was 12.9 months (3.5 and 50.0) in the AR group and 2.4 months (0.4 and 6.0) in the NR group (P=0.004). We examined whether the CTOT-04 signature score was associated with time from transplantation to biopsy and found the score was marginally associated (F-statistic: 0.3503, on 1 and 143 degrees of freedom, P=0.06328).^23^ We also evaluated whether the CTO-04 signature score was diagnostic of AR after controlling for time since transplantation to biopsy, and found it was diagnostic: P=3.88e-07 for the association without accounting for time since transplantation to biopsy compared to P=1.33e-06 including time in a general linear model.^24^

### CTOT-04 signature score discriminates kidney allograft recipients with NR biopsies from those with AR biopsies due to TCMR, ABMR or MR

The CTOT-04 signature, as expected from the earlier multicenter CTOT-04 study,^18^ was diagnostic of TCMR in the current investigation. Figure 2 shows the CTOT-04 signature score in each TCMR biopsy-matched urine sample. The median CTOT-04 signature score was -0.100 (-2.090 and 0.750) in the 47 urine samples matched to 47 TCMR biopsies and significantly higher than the score of -1.945 (-2.668 and -1.200) in the 50 urine samples matched to 50 NR biopsies (**Table 3**; P<0.0001, T test). At the CTOT-04 signature score cutpoint of -1.213, the signature distinguished urines matched to TCMR biopsies from urines matched to NR biopsies at a specificity of 76% and a sensitivity of 64% (**Table 4**; P<0.0001, Fischer’s Exact Test).

**Figure 2.**
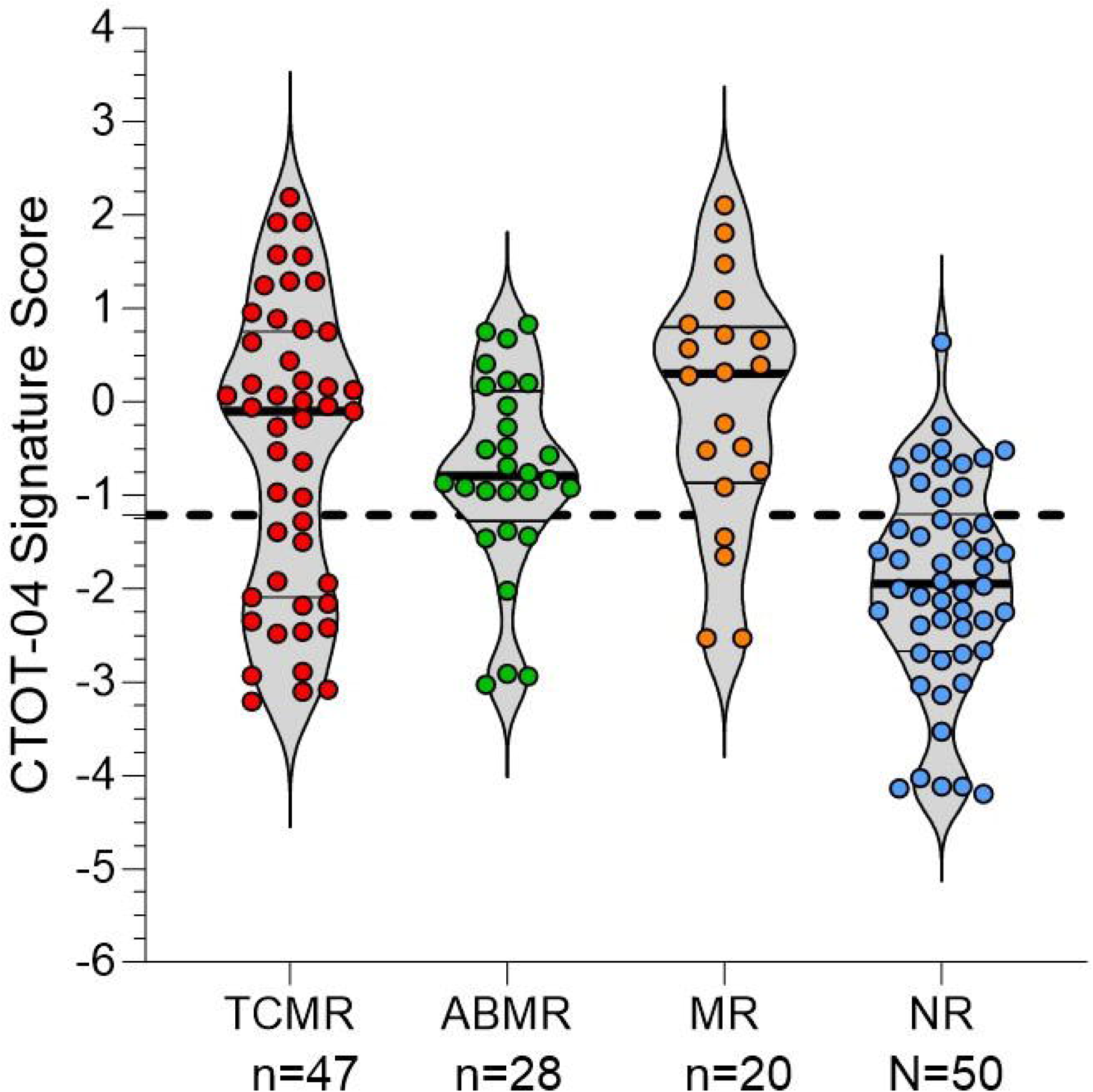
Violin Plots of CTOT-04 Signature Scores in Kidney Allograft Biopsy-Matched Urine Samples. Violin plots of CTOT-04 signature scores in the urine samples matched to TCMR biopsies (n=47), ABMR biopsies (n=28), Mixed Rejection (MR) biopsies (n=20), or NR biopsies (N=50). Customized RT-qPCR assays were used to measure urinary cell levels of CD3E mRNA, CXCL10 mRNA, and 18s rRNA in kidney allograft biopsy-matched urine samples collected from kidney allograft recipients and the CTOT-04 signature score was calculated from log_10_-transformed values for the 18S-normalized CD3E mRNA, 18S-normalized CXCL10 mRNA and 18S rRNA.^18^ The CTOT-04 signature score values of individual urine samples are shown as circles within the plot. The two thin black lines within each plot are the 25^th^ and 75^th^ percentile values, and thick black line is the 50^th^ (median) value of the CTOT-04 signature score. The horizontal dotted line parallel to the x-axis is the previously identified CTOT-04 signature score cutpoint of -1.213 that maximized sensitivity and specificity for the diagnosis of TCMR in the multicenter CTOT-04 study.^18^ One way analysis of variance comparing CTOT-04 signature score in urine samples matched to TCMR biopsies, ABMR biopsies, MR biopsies, or NR biopsies yielded a P value of <0.0001. Dunnett’s multiple comparison test for multiple group comparison yielded P <0.0001 for NR vs. TCMR; P <0.001 for NR vs. ABMR; and P <0.0001 for NR vs. MR.

The CTOT-04 signature score in each ABMR biopsy-matched urine sample is shown within the violin plot (Figure 2). The median CTOT-04 signature score in the 28 urine samples matched to 28 ABMR biopsies was -0.795 (-1.275 and 0.118) and significantly higher than the score of -1.945 (-2.668 and – 1.200) in the 50 urine samples matched to 50 NR biopsies (**Table 3**; P<0.001, T-test). At the CTOT-04 signature score cutpoint of -1.213, the signature distinguished urines matched to ABMR biopsies from urines matched to NR biopsies at a sensitivity of 75% and a specificity of 76% (**Table 4**; P<0.0001, Fischer’s Exact test).

The CTOT-04 signature score in each MR biopsy-matched urine sample is shown within the violin plot (Figure 2). The median CTOT-04 signature score in the 20 urines matched to 20 MR biopsies was 0.300 (-0.868 and 0.803) and significantly higher than the score of -1.945 (-2.668 and -1.200) in the 50 urines matched to 50 NR biopsies (**Table 3**, P<0.0001, T test). At the CTOT-04 signature score cutpoint of – 1.213, the signature distinguished urines matched to MR biopsies from urines matched to NR biopsies at a sensitivity of 80% and a specificity of 76% (**Table 4**; P<0.0001, Fischer’s Exact test).

The CTOT-04 signature score cutpoint of -1.213, developed for the diagnosis of TCMR,^18^ predicted TCMR, ABMR and MR in the current study with sensitivities and specificities that were not significantly different from the sensitivity and specificity for predicting TCMR in the CTOT-04 study (all comparisons, P>0.05).

### Diagnostic performance of CTOT-04 signature assessed using AUCs of ROC curves

We determined the diagnostic performance of the CTOT-04 signature across different types of AR biopsies by comparing AUCs of ROC curves. For robust statistical inference, we performed bootstrap replications by drawing random samples with replacements from the original data. We generated 5000 bootstrap sample sets and estimated the AUCs of ROC curves. The CTOT-04 signature discriminated TCMR biopsies from NR biopsies with an optimism-corrected AUC of 0.749 (95% CI, 0.638 to 0.840; P<0.0001)(Figure 3A); distinguished ABMR biopsies from NR biopsies with an optimism-corrected AUC of 0.780 (95% CI, 0.656 to 0.878; P<0.0001) (Figure 3B); discriminated MR biopsies from NR biopsies with an optimism-corrected AUC of 0.857 (Bias-corrected 95% CI, 0.727 to 0.947; P<0.0001) (Figure 3C); and distinguished AR biopsies from NR biopsies with an optimism-corrected AUC of 0.780 (95% CI, 0.700 to 0.846; P<0.0001) (Figure 3D).

**Figure 3.**
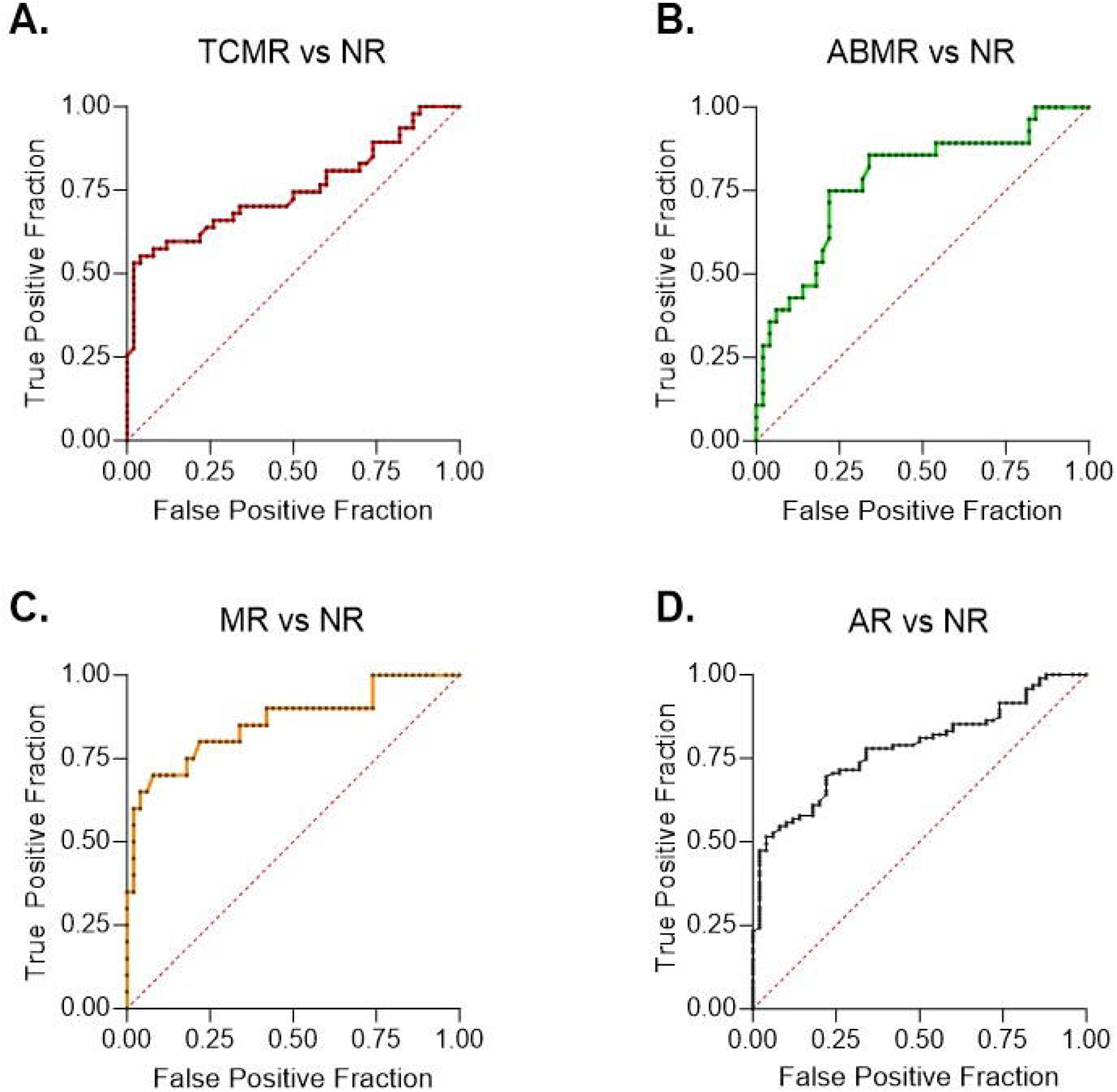
Receiver Operating Characteristic Curves For CTOT-04 Signature Scores in Kidney Allograft Biopsy-Matched Urine Samples. Receiver operating characteristic curve CTOT-04 signature scores in the urine samples matched to TCMR biopsies (A), ABMR biopsies (B), MR biopsies (C), or AR biopsies (D). The fraction of true positive results (Sensitivity) and the fraction of false positive results (1-Specificity) for the CTOT-04 signature score calculated from log_10_-transformed values for the 18S-normalized CD3E mRNA, 18S-normalized CXCL10 mRNA and 18S rRNA. A. In a comparison of patients with TCMR biopsies (n=47) with the patients with NR biopsies (n=50), the optimism-corrected AUC was 0.748 (Bias-corrected 95% CI, 0.638 to 0.840; P<0.0001). B. In a comparison of patients with ABMR biopsies (n=28) with the patients with NR biopsies (n=50), the optimism-corrected AUC was 0.780 (Bias-corrected 95% CI, 0.656 to 0.878; P<0.0001). C. In a comparison of patients with MR biopsies (n=20) with the patients with NR biopsies (n=50), the optimism-corrected AUC was 0.857 (Bias-corrected 95% CI, 0.727 to 0.947; P<0.0001). D. In a comparison of patients with AR biopsies (n=95) with the patients with NR biopsies (n=50), the optimism-corrected AUC was 0.780 (Bias-corrected 95% CI, 0.700 to 0.846; P<0.0001).

A comparison of the AUC of TCMR vs. NR to the AUC of ABMR vs. NR yielded a P value of 0.68; comparison of the AUC of TCMR vs. NR to the AUC of MR vs. NR yielded a P value of 0.15; and comparison of the AUC of TCMR vs. NR to the AUC of AR vs. NR yielded a P value of 0.61.

Altogether, our findings demonstrate that the CTOT-04 signature discriminates patients with NR biopsies from patients with AR, TCMR, ABMR or MR biopsies with similar ROC-AUC.

## DISCUSSION

In the current investigation, using an independent cohort of kidney allograft recipients, we validated that the urinary cell three-gene signature is diagnostic of TCMR. The optimism-corrected AUC of 0.75 (95% CI, 0.63 to 0.84) observed in the current study for discriminating TCMR biopsies from NR biopsies is almost identical to the AUC of 0.74 (95% CI, 0.61 to 0.86) for discriminating TCMR biopsies from NR biopsies in the validation set used in the CTOT-04 study.^18^ We further demonstrate that the three-gene signature is diagnostic of ABMR and MR with similar accuracy to that of TCMR. Altogether, our results suggest that the three-gene signature is diagnostic of all three major types of AR.

In the CTOT-04 study, there were only 10 biopsy-matched urine samples classified as ABMR and we did not analyze whether the signature is diagnostic of ABMR.^18^ Moreover, intragraft C4d was an essential criterion for ABMR and chronic active ABMR was not an entity in the Banff schema^26^ used to classify biopsies in the CTOT-04 study. In this study, we analyzed a larger sample size – 48 urine samples matched to biopsies showing either ABMR alone or with concurrent TCMR. Moreover, we captured the broad spectrum of ABMR by including urine samples matched to active ABMR, chronic active ABMR or concurrent ABMR+TCMR, and used 2019 Banff criteria for ABMR.^4^ The Banff classification schema is an ever-evolving process and challenges exist in biopsy diagnosis of chronic active ABMR.^27^ Notwithstanding these considerations, our new observations broaden the diagnostic scope of the CTOT-04 signature from predicting acute rejection due to TCMR to predicting acute rejection due to TCMR, ABMR or MR. This is a significant advantage since it is difficult to identify a priori the type of acute rejection in a biopsy based on clinical parameters such as increase in serum creatinine.

Our formulation and testing of the hypothesis that the CTOT-04 signature would serve as a noninvasive biomarker of all three major types of acute rejection were stimulated in part by our findings from RNA sequencing of kidney allografts and biopsy-matched urines.^19^ The pertinent findings include differential gene expression (DEG) analysis identifying shared gene sets and enriched KEGG gene pathways^28^ in urine matched to TCMR biopsies and urine matched to ABMR biopsies. Using stringent criteria (Log_2_ fold-change>2 and FDR<0.05) for DEG, we found 127 genes shared between urine matched to TCMR biopsies and urine matched to ABMR biopsies.^19^ Furthermore, gene set enrichment analysis^29^ identified that DEGs in TCMR biopsies vs. NR biopsies are not only enriched in TCMR biopsy-matched urines but also in ABMR biopsy-matched urines, and DEGs in ABMR biopsies vs. NR biopsies are enriched in both ABMR biopsy-matched urines and TCMR biopsy-matched urines.^19^

The biological activities of the proteins encoded by the mRNAs included in the parsimonious CTOT-04 signature-CD3E mRNA and CXCL10 mRNA-likely contributed to the signature being informative of all three types of acute rejection. CD3E protein is an integral component of T cell antigen recognition complex and essential for transmembrane signaling of T cells. T cells play a non-redundant role in T cell-mediated rejection and multiple T subtypes have been identified in the human kidney allograft undergoing TCMR. T follicular helper cells aid in the B cell production of antibodies including anti-HLA antibodies, the primary mediators of ABMR. In our transcriptomic profiling of kidney allografts and urinary cells, the abundance of CD3E mRNA was higher in urine matched to TCMR biopsies vs. urine matched to NR biopsies and in urine matched to ABMR biopsies vs. urine matched to NR biopsies. CD3E mRNA was among the enriched mRNAs shared between urine matched to TCMR biopsies and urine matched to ABMR biopsies.^19^

CXCL10 mRNA was the other component of the CTOT-04 signature developed to diagnose TCMR. Both the mRNA and the encoded chemoattractant have been linked to ABMR in multiple studies.^16, 30–32^ In our unbiassed whole transcriptome profiling of kidney allograft biopsies and urinary cells, CXCL10 transcripts were higher in urine matched to TCMR biopsies vs. urine matched to NR biopsies and in urine matched to ABMR biopsies vs. urine matched to NR biopsies. CXCL10 mRNA was among the enriched mRNAs shared between urine matched to TCMR biopsies and urine matched to ABMR biopsies.^19^

The clinical significance of the CTOT-04 signature being diagnostic of TCMR, ABMR or MR is at least two-fold. First, the CTOT-04 signature could function as a surrogate for the invasive biopsy performed to distinguish kidney allograft recipients with AR due to TCMR, ABMR, or MR from recipients without rejection. Second, the findings that CD3E mRNA and CXCL10 mRNA are overexpressed in urine matched to TCMR, ABMR or MR biopsies suggest the hypothesis that these mRNAs or the proteins they encode might serve as mechanism-based targets to treat acute rejection due to all three major subtypes.

This is especially important for ABMR and MR since there are no effective therapies for acute rejection mediated by antibodies.^9^

Our study has limitations. This is a single-center study; a multicenter study, fashioned after the CTOT-04 investigation, is worthy of consideration. Although we have validated the three-gene signature as being diagnostic of AR in each instance by biopsy diagnosis and by bootstrap resampling, validation using an external cohort may be of value and the proposed multicenter study would also serve this purpose.

Altogether, we demonstrate that the CTOT-04 three-gene signature is not only diagnostic of TCMR in kidney allografts but is also predictive of ABMR and MR with performance characteristics similar to those of TCMR. Our development of a noninvasive and potentially mechanistic signature of acute rejection due to TCMR, ABMR or MR could reduce the need for the invasive biopsy procedure as well as suggest novel therapeutic targets for the treatment of acute rejection.

## Data Availability

All data produced in the present study are available upon reasonable request to the authors

## Acknowledgments

The authors gratefully acknowledge the exceptional contribution of our colleagues, Dr. Steven Salvatore, Dr. Surya V. Seshan, Dr. Michelle Lubetzky, Dr. Ruchuang Ding, Shady Albakry, Michael Cassidy, Rex Friedlander, and Gabriel Stryjniak. We thank our dedicated NewYork Presbyterian-Weill Cornell Medicine kidney transplant team and particularly our patients.

## Declaration of generative AI in scientific writing

The authors declare that generative artificial intelligence (AI) or AI-assisted technologies were not used in the preparation of this manuscript and the writing was done exclusively by the authors.

